# Explainable machine learning to identify patients at risk of developing hospital acquired infections

**DOI:** 10.1101/2024.11.13.24317108

**Authors:** Andrew P. Creagh, Tom Pease, Philip Ashworth, Lloyd Bradley, Sophie Duport

**Affiliations:** Sanome, London, UK; PatientSource, Cambridge, UK; Royal Hospital for Neuro-disability, London, UK

**Keywords:** clinical machine learning, infection risk stratification, clinical decision support tools, specialised neurological conditions, electronic health records

## Abstract

Hospital-acquired infections (HAIs) contribute to increased mortality rates and extended hospital stays. Patients with complex neurological impairments, secondary to conditions such as acquired brain injury or progressive degenerative conditions are particularly prone to HAIs and often have the worst resulting clinical outcomes and highest associated cost of care. Research indicates that the prompt identification of such infections can significantly mitigate mortality rates and reduce hospitalisation duration. The current standard of care for timely detection of HAIs for inpatient acute and post-acute care settings in the UK is the National Early Warning Score v02 (NEWS2). NEWS2, despite its strengths, has been shown to have poor prognostic accuracy for specific indications, such as infections. This study developed a machine learning (ML) based risk stratification tool, utilising routinely collected patient electronic health record (EHR) data, encompassing over 800+ patients and 400k+ observations collected across 4-years, aimed at predicting the likelihood of infection in patients within an inpatient care setting for patients with complex acquired neurological conditions. Built with a combination of historical patient data, clinical coding, observations, clinician reported outcomes, and textual data, we evaluated our framework to identify individuals with an elevated risk of infection within a 7-day time-frame, retrospectively over a 1-year “silent-mode” evaluation. We investigated several time-to-event model configurations, including manual feature-based and data-driven deep generative techniques, to jointly estimate the timing and risk of infection onset. We observed strong performance of the models developed in this study, achieving high prognostic accuracy and robust calibration from 72–6 hours prior to clinical suspicion of infection, with AUROC values ranging from 0.776–0.889 and well-calibrated risk estimates exhibited across those time intervals (IBS<0.178). Furthermore, by assigning model-generated risk scores into distinct categories (low, moderate, high, severe), we effectively stratified patients with a higher susceptibility to infections from those with lower risk profiles. Post-hoc explainability analysis provided valuable insights into key risk factors, such as vital signs, recent infection history, and patient age, which aligned well with prior clinical knowledge. Our findings highlight our framework’s potential for accurate and explainable insights, facilitating clinician trust and supporting integration into real-world patient care workflows. Given the heterogeneous and complex patient population, and our under-utilisation of the data recorded in routine clinical notes and lab reports, there are considerable opportunities for performance improvement in future research by expanding our model’s multimodal capabilities, generalisability, and additional model personalisation steps.

## 1 Introduction

Hospital-acquired infections (HAI), also referred to as healthcare-associated infections, are infections which are not present at the time of admission. They are typically classified as infections diagnosed at least 48-hours after hospital admission [1]. HAIs have long been recognised as one of the most significant burdens on global healthcare systems [2], and there is a substantial body of evidence analysing the prevalence of and morbidity associated with HAIs [3], with some studies suggesting that HAIs are the leading cause of mortality and morbidity worldwide [4].

The current standard of care for the timely detection of HAIs in UK hospitals are general clinical deterioration rule-based Early Warning Scores (EWS) such as National Early Warning Score v02 (NEWS2) [5] and Modified Early Warning Score (MEWS) [6]. Machine learning (ML) algorithms present a clear opportunity to identify HAIs earlier and with better accuracy than these manual rules-based methods. For instance, models can be trained to predict HAIs specifically, and state-of-the-art machine learning algorithms are shown to outperform rules-based standards of care, such as NEWS2[7]–[9] and MEWS [10]–[12]. This is likely to do with a combination of the wider range of data points used by the ML models, as well as their ability to capture non-linearities in the data.

Effective ML EWS could improve patient outcomes or reduce healthcare costs by alleviating pressure on services, for example, by reducing the likelihood of readmissions [13]. Early warning of HAI could also indirectly decrease the likelihood of developing other healthcare-associated conditions, or antibiotic resistance through more targeted treatment [14]. For instance, preventing avoidable HAIs could save NHS England £410M (or £2.7M per trust) through 10% reduced length-of-stay and increased patient flow alone [15]–[17].

Despite the known prevalence and burden of HAIs, there are relatively few examples of solutions that develop ML EWS for HAIs in general [18]–[20], or indeed for other conditions such as urinary tract infection [21]–[23], clostridium difficile infection [24], [25] and central line-associated bloodstream infection[26]. These papers do, however, demonstrate the feasibility of using ML to predict HAIs in clinical settings, with reported area under the receiver operator curve (AUROC) scores ranging from 0.746 to 0.884. Most studies train and evaluate models to produce a single prediction at a fixed point in each patient’s care episode [22], [23], [25], [26], but there are also several examples of studies which instead train and evaluate models over a fixed time-horizon (typically hours) before an infection event [18]–[20]. None of the papers described algorithms which had been deployed and used in live medical settings, nor have any papers investigated models specifically for patients with complex neurological impairments, as this paper will go on to do.

Whilst the literature on EWS for HAIs is still developing, there is a much more extensive literature on relevant related tasks: ML EWS for both sepsis and general deterioration, the majority in intensive care unit (ICU) settings. The plethora of retrospective studies [13], [27], and deployed ML EWS [28], [29] confirm viability of using ML to create models with strong predictive performance, even in the context of general acute hospital wards [30]–[32] where data is significantly less granular and detailed than in an ICU. Studies draw on a large range of algorithmic approaches to suit the needs of their task and their objectives, including traditional linear modes [9], [33], decision tree based models [34], [35], deep learning approaches [36], [37], and survival analysis based models [38], [39]. Depending on the goals of the application and the data available in the setting, projects vary hugely in their complexity, from model pipelines with just a few variables [37] to hundreds [40]. Finally it is interesting to note that, despite the longitudinal nature of the data available, and the fact that predicting deterioration is a time-sensitive problem, most papers do not elect to use explicit time-series approaches. Instead, it is common to combine the use of statistical models, for their ease of implementation and interpretability, with feature engineering to create ‘look-back features’ which give models access to information about a patient’s recent history. This approach can vary in complexity from a few summary features like the mean, minimum and maximum of very recent data [41], to generating hundreds of features from more detailed statistics about a patient’s hospital stay [42]. Noteworthy, the latter paper, by Hyland et al. (2020), has shown state-of-the-art performance of careful feature engineering in comparison to deep learning models investigated.

In this study, we build on the current state of the field by developing a machine learning-based clinical decision support (CDS) tool to estimate a patient’s risk of developing nosocomial infections in an inpatient care setting for patients with complex acquired neurological conditions. Patients with complex neurological impairments secondary to conditions such as acquired brain injury or progressive degenerative conditions are highly susceptible to healthcare associated infections, for a range of reasons including possible immune dysfunction [43]. For example, over 15% of severe acquired brain injury patients develop a HAI, with mortality rates of 20-30% [44]. HAIs in these patients lead to poorer outcomes [45], [46], longer stays [47], development of antibiotic resistance of broad-spectrum antibiotics for prophylactic and therapeutic purposes [48]. Furthermore, due to communication difficulties, cognitive impairment, and atypical baseline vital signs in patients with complex neurological conditions [49], tools like NEWS2 may be less effective and HAIs harder to diagnose.

Here, we present our ML framework built using large-scale retrospective data, aggregating readily available patient electronic health record (EHR) data (collected on 800+ patients, 400k+ assessments) with patient medical history and recent trends. This approach creates personalised risk prediction models that are inherently explainable and understandable, where predictions are displayed back into the EHR to support clinical decision making by healthcare professionals. We compare several state-of-the-art methodologies to develop survival task heads, across both manually extracted and automatically extracted features, as learned via deep generative techniques. Finally, through “silent-mode” retrospective evaluation, we explore each model’s performance to the established clinical scoring systems, such as NEWS2, which hypothesises model performance if it were to be deployed in clinical practice (baseline risk) in a real-world prospective validation.

## 2 Methodology

### 2.1 Dataset

In this study, longitudinal data was collected on patients with complex neurological disabilities. Retrospective data (800+ patients, 400K+ observations) was obtained over 4-year period (Jul. 2019–Aug. 2023) at a co-pilot site, the Royal Hospital for Neuro-Disability (RHN), London, UK. This study was conducted in accordance with the ethical principles outlined in the Declaration of Helsinki and received favourable ethical opinion from NHS Research Ethics Committee 24/NE/0008 supported by the Confidentiality Advisory Group 23/CAG/0110. All data handling and analysis were performed in compliance with confidentiality guidelines and data protection regulations. The use of retrospective patient data was ethically reviewed, ensuring the study adhered to principles of transparency, data privacy, and minimal risk to participants.

Electronic health record (EHR) data obtained via PatientSource, UK, consisting of meta-data, such as patient demographics (age, sex, etc.), as well as physical measurements, such as height, weight, etc., were collected along with raw time-series observations, e.g. heart rate, temperature, etc., and clinical reported outcomes, e.g., Glasgow Coma Scale (GCS). Patient co-morbidities and histories were obtained from EHR coding, for example ICD-10 coding, as well as from patient clinical notes and ward information. The study population consists of adult in-patients (aged ≥18 and <80 years) admitted to RHN during the recruitment period, specifically those diagnosed with brain injuries, neurological conditions, or degenerative/progressive disorders, provided they do not meet any exclusion criteria. Exclusion criteria included patients under 18 years of age, those who develop an infection within 48-hours of initial admission, and those for whom all clinical observations fall under exclusion parameters. Additional exclusions were applied to patients actively participating in, or recently involved (within the last 30 days) in, interventional clinical trials that might influence the algorithm developed in this work, pregnant patients, those with an imminent and inevitable prognosis of death as determined by the investigator, and patients who were previously enrolled in the study.

We follow a retrospective validation approach, which mimics the design of a year-long blinded observational cohort study. This design enabled a determination of model performance—if the model were to be deployed— in comparison to standard-of-care. This methodology of temporal validation is comparable to retrospective “silent-mode” validation [50]–[52]. To create the development (in-sample training) and validation (held-out testing) datasets, we utilised temporal validation with a 80:20 split (i.e., time-series cross validation with a held-out set), which ensured that the validation process respects the temporal order of data. The dataset set for development and validation contained data based at the same site, however, the development dataset is historic and the validation dataset contains recent patient data. The training (Jul. 2019–Sep. 2022; 80%, ≈1200 days) and held-out testing (Oct. 2022–Aug. 2023; 20% ≈300 days) sets were independent and there are no patients who are present in both development and validation sets and a 3-week gap was excluded from the end of each train set before the test set. Fig. 1 details the study design and evaluation. The population distribution differences between development and validation sets are presented in Table 1.

**Table 1:** Summary of Population Characteristics. The median + 95^th^ centile of each characteristic are reported across the entire population, the historical in-sample training split, and the temporally held-out test split (1-year silent-mode), unless otherwise stated.

|  | population | historical training set | held-out testing set |
| --- | --- | --- | --- |
| **Data Collection** |  |  |  |
| Timeline, year |  | 2019–2022 | 2022–2023 |
| period, weeks (%) |  | 170 (80%) | 45 (20%) |
| patients, n (%) | 824 | 692 (84%) | 128 (16%) |
| assessments, n (%) | 408k | 386k (95%) | 22k (5%) |
| observations, n (%) | 4.89M | 4.63M (92%) | 264k (5%) |
| **Demographics** |  |  |  |
| Age, years | 53 (23–76) | 53 (24–76) | 56 (27–75) |
| Female, n (%) | 317 (38%) | 276 (40%) | 40 (31%) |
| **Patient History<sup>1</sup>** |  |  |  |
| Disease Status, n (%) |  |  |  |
| cardiovascular disease | 119 (14%) | 98 (14%) | 21 (16%) |
| endocrine disorder | 146 (18%) | 118 (17%) | 28 (22%) |
| epilepsy | 78 (9%) | 72 (10%) | 6 (5%) |
| musculoskeletal disorder | 75 (9%) | 64 (9%) | 11 (9%) |
| neurological disorder | 384 (47%) | 323 (47%) | 61 (48%) |
| respiratory disease | 126 (15%) | 109 (16%) | 17 (13%) |
| vascular disease | 182 (22%) | 147 (21%) | 35 (27%) |
| Other comorbidity, n (%) |  |  |  |
| diffuse brain injury | 73 (9%) | 59 (9%) | 14 (11%) |
| hypertension | 167 (20%) | 134 (19%) | 33 (26%) |
| brain hypoxia-anoxia | 43 (5%) | 29 (4%) | 14 (11%) |
| subarachnoid haemorrhage | 83 (10%) | 63 (9%) | 20 (16%) |
| haemorrhagic stroke | 30 (4%) | 18 (3%) | 12 (9%) |
| stroke (non-specified) | 46 (6%) | 34 (5%) | 12 (9%) |
| gastrostomy-peg | 100 (12%) | 94 (14%) | 6 (5%) |
| type-2 diabetes | 103 (12%) | 82 (12%) | 21 (16%) |
| prev. surgery (non-specific) | 81 (10%) | 74 (11%) | 7 (5%) |
| prev. surgery (neurosurgical) | 64 (8%) | 58 (8%) | 5 (4%) |
| prev. surgery (orthopaedic) | 66 (8%) | 57 (8%) | 9 (7%) |
| prev. cardiac arrest | 53 (6%) | 39 (6%) | 14 (11%) |
| **Study characteristic(s)** |  |  |  |
| Patient cohort(s)<sup>2</sup>, n (%) |  |  |  |
| Brain Injury | 166 (15%) | 129 (14%) | 37 (20%) |
| Long-term Stay | 62 (5%) | 58 (6%) | 4 (2%) |
| Neurological Disorder | 94 (8%) | 81 (8%) | 13 (7%) |
| Other<sup>a</sup> | 131 (11%) | 110 (12%) | 21 (11%) |
| PDoC<sup>b</sup> | 215 (19%) | 194 (20%) | 21 (11%) |
| Rehab | 259 (23%) | 228 (24%) | 31 (16%) |
| Stroke | 215 (19%) | 153 (16%) | 62 (33%) |
| Enrolment, months | 20 (2–170) | 23 (3–170) | 14 (1–39) |
| Observations, per day<sup>c</sup> | 1 (2–21) | 1 (2–16) | 1 (2–14) |
| Infections<sup>3</sup>, n (%) |  |  |  |
| total | 2355 (0.9%) | 2132 (0.9%) | 223 (1.1%) |
| per-patient | 2 (1–9) | 2 (1–9) | 1 (1–4) |
| censoring<sup>d</sup>, % | 93% | 92% | 90% |
| Incomplete assessments, n (%) | 7595 (3%) | 7087 (3%) | 508 (2.5%) |

<sup>1</sup> Most common disease/disorder-type and comorbidity observed, loosely grouped.
<sup>2</sup> Note, patients may change characteristic or be denoted by multiple characteristics over data collection period.
<sup>3</sup> Infections are defined by the timing of acute antibiotic prescriptions (non-prophylactically administered, non-topically administered antibiotics).
<sup>a</sup> Patients with other neuro-disability and/or neuro-behavioural conditions;
<sup>b</sup> PDoC, Patients with Prolonged Disorders of Consciousness;
<sup>c</sup> Median (Min-Max) values reported.
<sup>d</sup> Right-censoring, 7-days from measurement collection.

**Figure 1:**
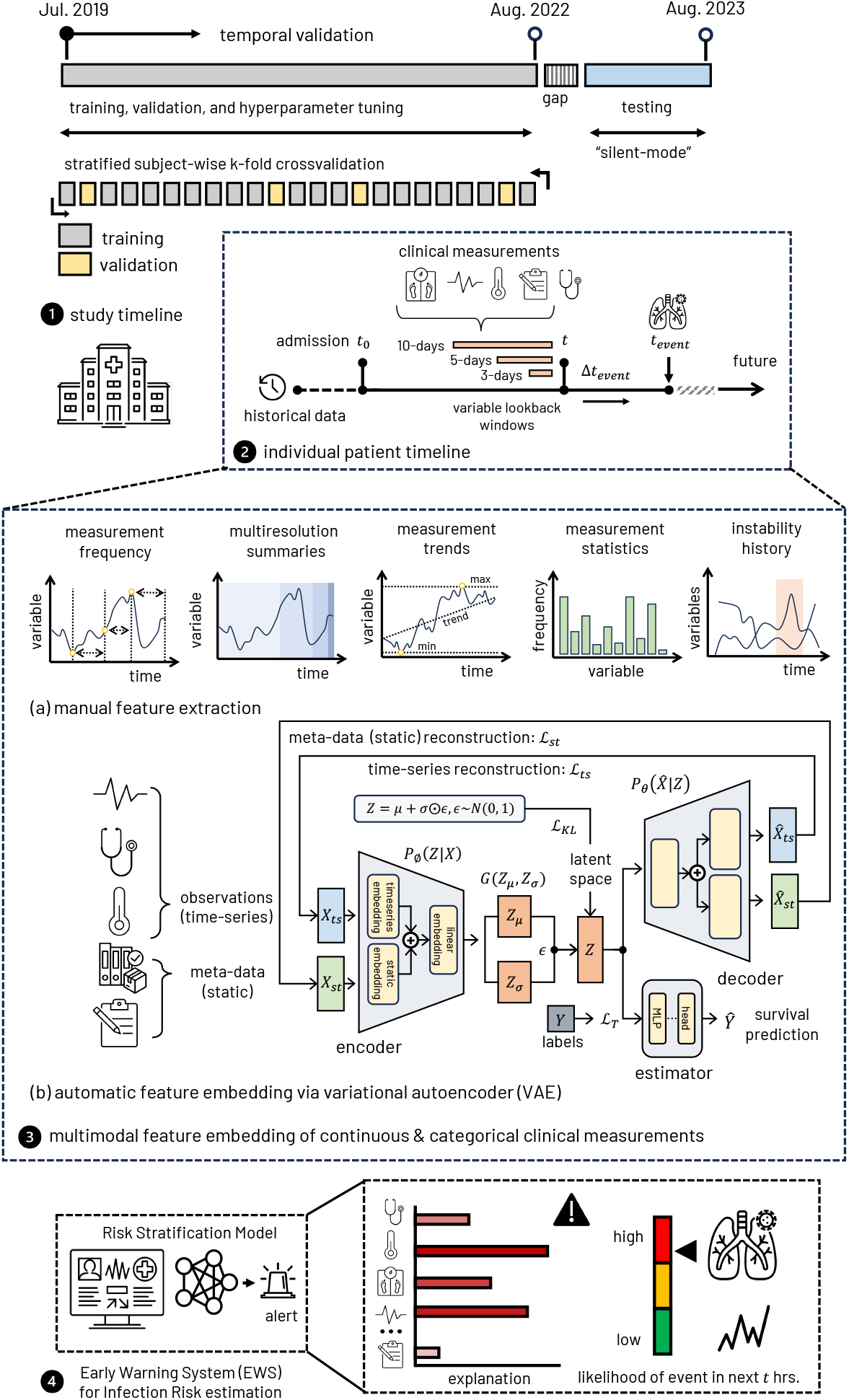
Overview of study design, feature extraction, and infection risk estimation workflow. Retrospective data collection timeline spans from Jul. 2019–Aug. 2023. Training, validation, and hyperparameter tuning follows a stratified subject-wise k-fold cross-validation framework. Temporal validation enables a “silent-mode” testing phase, which mimics the design of a year-long blinded study. (panel 1). Individual patient timelines combined patient history with clinical measurements collected from admission (*t*_0_) to infection events (*t*_*event*_) (panel 2). Manual feature extraction (panel 3a) with variable lookback windows included summaries of measurement frequency, trends, and instability, while automatic feature embedding via a variational autoencoder (VAE) generated data-driven features (panel 3b). These data respresentations were utalised in the infection risk stratification model and integrated as an early warning system (EWS) that continuously stratifies infection risk levels over a fixed 4time window, *t* in the future (panel 4).

### 2.2 Processing

Data was processed following a number of steps: wrangling and collating EHR data per patient, per visit ID; collating static meta-data, such as demographics and covariates obtained from ICD-10 codes; concatenating anthropometric measurements, such as height, weight, BMI; temporally aligning raw time-series observations, e.g. heart rate, temperature, etc., as well as infrequently captured data clinical reported outcomes, e.g., Glasgow Coma Scale (GCS), and textual features manually extracted from the clinical notes (for example, persistent coughing). Historical data from patient histories was also extracted and combined, such as the number of previous events, the time since last event or time-differences between concurrent observations. Manual outlier removal was performed, removing implausible or out-of-clinical-range values for data with known ranges or categories.

Summary features and statistics were also calculated across the raw observations over multi-resolution summaries look-back windows of varying lengths [*t* = 3, 5, 10] days, following procedures introduced by [42]. See figure 1 for further details.

Missing observational data were first imputed using the “carry-last-value-forward” method for each patient sequence up to a maximum of 48 hours. Non-carried missing values were imputed via single imputation using the mean values in the training data for numeric data and the most frequent category for categorical data. Numeric data was standardised using by removing the mean and scaling to unit variance, based on values obtained from the training set. Categorical hand-crafted features were normalised in an analogous step using categorical boosting (catboost), a variation of target encoding [53].

Infection event annotation, i.e. “labelling”, indicating clinical suspicion of infection was determined through timestamped confirmed antibiotic prescription and/or administration of an antibiotic associated with infection; similar approaches are taken when standard hospital coding indicating the timing of an infection were unavailable or unreliable [18], [19]. As such, we used the earliest timing of the antibiotic orders to mark clinical suspicion of infection for the given patient. Events associated with prophylactic antibiotics were removed from the dataset. In order to infer that each infection is “hospital-acquired” or “nosocomial”, data from the first 48-hours after admission were removed from the dataset. Observations following a 14-day buffer period after first administration of an antiobiotic were removed from the dataset, ensuring only the onset of infection was targeted. For every patient sequence, we calculated the duration (i.e., Δ*t*) to the next relevant patient antibiotic label, in chronological order, as such denoting the future time-to-event, 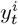 for each observation set; Δ*t*^*i*^, for patients with no next relevant event were masked using the mean timedelta of the training set.

### 2.3 Model

We formulated our task as time-to-event failure analysis problem, incorporating dynamic survival analysis models, to jointly model time and the likelihood of an event by estimating the survival function, *S*(*t*). The survival function *S*(*t*) returns the probability of survival beyond time *t*:

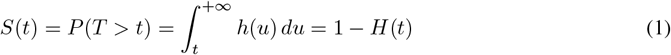

where *H*(*t*) is the the cumulative hazard function, and the hazard function *h*(*t*) denotes an approximate probability (it is not bounded from above) that an event occurs in the small time interval [*t*; *t* + Δ*t*], under the condition that an individual would remain event-free up to time *t*:

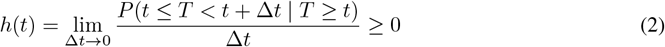

The model-estimated survival function for an individual patient with data **x**, defined as:

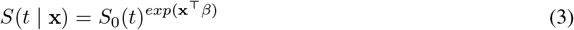

where *S*_0_(*t*) is the baseline survival function, estimated by Breslow’s estimator.

We formulated the task as a time-to-event analysis with a right-censoring window of *T* = 8 days. Observations with *t* ≤7 days were considered positive infection events, assigned a time-to-event (Δ*t*) in days, recorded with minute precision. Observations with *t >* 7 days were right-censored at *t* = 8 days. This threshold was selected to capture the typical range of incubation periods from exposure to symptom onset for many infections [54].

We compared popular survival analysis task heads, spanning linear and non-linear methodologies, the standard Cox-Proportional hazards model (CoxPH) extended with an elastic net penalty [55], to Gradient Boosted Survival Trees (GboostSurv) [56]–[58] (with the partial likelihood of the proportional hazards model as the objective function [59]), as well as discretized-time models, such as Logistic-Hazard (LogHz) [60] (also known as Nnet-survival [61]) and DeepHit [62]. We experimented with parametrising these models using the raw feature data, as well as incorporating data-driven (generative) feature embedding via a variational auto-encoder (VAE).

Our conditional VAE architecture incorporated multiple layers of Long Short-Term Memory (LSTM) networks to model both static and time-varying features for downstream survival analysis (i.e. infection risk) [63], [64]. The encoder processes static covariates—including raw categorical variables transformed via entity embeddings—and sequential numeric observations, encoding them into a latent space represented by a multivariate Gaussian distribution with mean vector *µ* and standard deviation vector *σ*. The LSTM-based decoder, reconstructs the features from the latent representation. The use of entity embeddings for categorical variables allows the model to convert discrete features into continuous representations, facilitating their integration into the latent space. Additionally, the LSTM networks within the VAE framework enable the model to capture temporal dependencies inherent in our sequential data (e.g., vitals, or other observations). Similarly to [65], in addition to the encoder-decoder structure, our VAE model was conditioned to jointly learn a survival task head to predict time-to-event for our infection estimation task.

We experimented with optimizing over multiple VAE loss terms: the reconstruction losses (ℒ_*ts*_ and ℒ_*st*_), calculated as the mean squared error (MSE) and cross entropy (CE) loss between the original numeric (time-series) and (static) categorical inputs and the reconstructed output, respectively, the prediction loss (ℒ_*T*_) —either the partial likelihood of the proportional hazards [59] or bespoke DeepHit [62] or LogHz [61] loss functions—as well as the Kullback–Leibler (KL) divergence (ℒ_*KL*_), which measures the difference between the learned latent distribution *q*(**z** | **x**) and a prior multivariate Gaussian distribution *p*(**z**). The reparameterisation trick is employed to enable backpropagation through the stochastic sampling process by expressing the sampled latent vector **z** as **z** = *µ* + *σ* ⊙ ***ϵ***, where ***ϵ*** is sampled from a standard normal distribution 𝒩 (**0, I**), and ⊙denotes element-wise multiplication. By minimising a combined weighted loss function, the model aims to accurately reconstruct input data while distilling the latent space to capture the underlying data distribution. The latent representation **z** effectively captures both static and dynamic aspects of the data, providing a feature space suitable for downstream survival analysis tasks.

### 2.4 Model Training

For each model developed, hyperparameter values were determined through stratified patient-wise (i.e., the data is stratified on the basis of patient, meaning that no patient contributes observations to both the training and validation set) 5-fold cross-validation and randomised grid search using the respective training sets with 50–100 candidate parameter combinations, depending on the model and its structure. This approach ensured that different combinations of hyperparameter values were evaluated on as much data as possible to provide the best estimate of model performance on unseen data, allowing us to determine the optimal settings for model training. We also experimented with optimising over different scoring metrics: *C*_IPCW_, 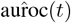, and the Integrated Brier Score (IBS).

Regularisation was performed via data augmentation independently for time series variables (e.g., vitals) as described by [66], and for categorical variables following approaches introduced by [67]. A weighted oversampling scheme was applied during training for class rebalancing caused by the high right-censoring observed in the data. Evaluation of models maintained the original right-censored distribution.

Models were trained for 50 epochs with early stopping implemented if the validation loss did not improve after 5 consecutive epochs. Additionally, a 3-epoch warm-up period was used at the beginning of training to stabilise the learning process.

We employed a linear scaling of the learning rate, adjusting it according to the batch size to maintain consistent training dynamics. Furthermore, we utilised a secondary learning rate reduction on plateau strategy, where the learning rate was reduced when the validation loss plateaued, to facilitate better convergence. To enhance the training process, we utilised teacher forcing during decoder training and applied Kullback–Leibler (KL) divergence annealing to balance the reconstruction and regularisation terms in the loss function. Gradient clipping was implemented to prevent exploding gradients and ensure numerical stability during training.

### 2.5 Model Evaluation

#### 2.5.1 Metrics

We assessed model performance using survival-specific metrics, which account for right-censored data and time-to-event outcome, the concordance index (C-index), time-dependent area under the receiver operating characteristic curve (time-dependent AUROC), and the integrated Brier score (IBS). Additional visualisation of outcome evaluation was also performed using the Kaplan-Meier Estimator. In order to determine patient generalisbility of our model predictions, metrics were calculated over 100 random bootstrap patient samples of the out-of-sample data and reported as the median+IQR or median+95^th^ centile, where applicable.

##### Concordance index (C-index)

The C-index quantifies the model’s proficiency in correctly ordering pairs of subjects with respect to their times to event:

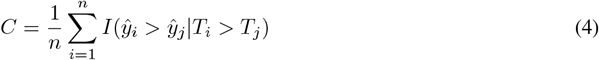

Here, *T*_*i*_ and *T*_*j*_ represent the observed event times for any two subjects, while *ŷ*_*i*_ and *ŷ*_*j*_ denote the corresponding predicted survival times or risks. However, a notable limitation is its aggregate nature, which overlooks the precise timing of events, focusing instead on ordinal rankings. Given the high-prominence of right-censoring in our dataset, we have used the adapted version of the C-index, which incorporates the inverse probability of censoring weights (IPCW). The IPCW C-index adjustment methodologically recalibrates the contributions of each subject pair in the C-index calculation, employing weights (*w*_*i*_) that inversely correspond to their probability of censorship. The IPCW-adjusted C-index, denoted as *C*_*IP CW*_, is mathematically expressed as

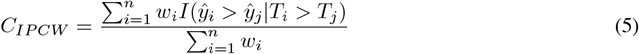

where *w*_*i*_ represents the censoring weight for the *i*^*th*^ subject, ensuring that the metric accurately reflects the model’s predictive performance by mitigating the bias towards subjects with observed events.

##### Time-dependent area under the receiver operating characteristic curve

The receiver operating characteristic (ROC) curve and the area under the ROC curve (AUROC) can be extended to survival data by defining sensitivity (true positive rate) and specificity (true negative rate) as time-dependent measures. This adaptation facilitates a more granular evaluation of model performance, particularly in distinguishing between subjects who experience events and those who do not at various points in time. *Cumulative cases* are all individuals that experienced an event prior to or at time *t* (*t*_*i*_ ≤*t*), whereas *dynamic controls* are those with *t*_*i*_ *> t*. The associated cumulative/dynamic auroc quantifies how well a model can distinguish subjects who fail by a given time (*t*_*i*_ ≤*t*) from subjects who fail after this time (*t*_*i*_ *> t*). Given an estimator of the *i*-th individual’s risk score 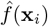, the cumulative/dynamic AUC at time *t* is defined as

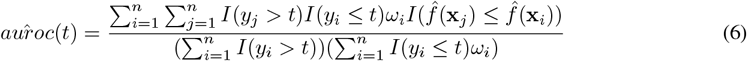

where *ω*_*i*_ are inverse probability of censoring weights (IPCW).

##### Integrated Brier Score

The time-dependent Brier score is the mean squared error at time point *t*:

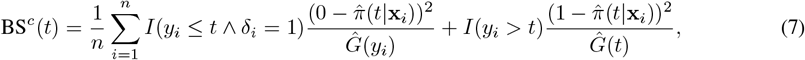

where 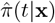 is the predicted probability of remaining event-free up to time point *t* for a feature vector **x**, and 1*/Ĝ*(*t*) is an inverse probability of censoring weight, estimated by the Kaplan-Meier estimator. The IBS encapsulates an assessment of a model’s calibration and discrimination prowess over the entirety of the time-horizon period, *t*_1_ *≤ t ≤ t*_max_, by integrating the time-dependent Brier score over the interval [*t*_1_; *t*_max_]:

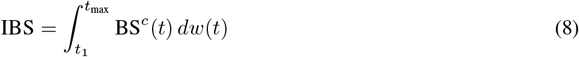

where the weighting function is *w*(*t*) = *t/t*_max_. The integral is estimated via the trapezoidal rule.

#### 2.5.2 Model Interpretation

Model-estimated posterior risk scores were segmented into ordinal risk-levels, [low, moderate, high, or severe]-risk. Thresholds for risk-levels were determined by discretising the distribution of in-sample risk-scores [log-hazard-ratios]. We experimented with fixed interval and quartiles thresholds. Thresholds were then applied to the held-out test data to determine model-estimated risk-levels.

Post-hoc model explainability was determined using SHapley Additive exPlanations (SHAP) values, which were utilised to further understand the contribution of individual features to the predictive models, helping to understand feature importance and model performance [68]. This approach helps to facilitate the interpretation of complex survival models at the individual prediction level, but also the relative importance of features across the model.

#### 2.5.3 Statistical Analysis

The non-parametric log-rank test used to compare the survival distributions of two samples [69], [70], with null hypothesis that there was no difference in the survival distributions between two samples. Differences between survival outcomes metrics for models and NEWS2 outcomes were assessed with the non-parametric Wilcoxon Signed-Rank test [71], comparing two related samples or repeated measurements on a single sample to assess whether their population median ranks differed. Simple sample size calculation using two proportions was used to estimate the required number of sample bootstraps needed to evaluate per-event metrics (e.g. c-index, auroc, IBS). To compute the required sample size needed per group (and the subsequent total number of events required) to determine statistical significance for survival-based outcomes (e.g., outcomes from the log-rank test), we applied a censoring-adjusted sample size calculation, calculated empirically in the training dataset [72], [73]. All p-values are corrected for multiple hypothesis testing (i.e., using multiple evaluation metrics) using using the linear step-up procedure introduced by Benjamini and Hochberg (BH) [74].

## 3 Results

### 3.1 Model comparison against standard-of-care

We developed multiple pipeline configurations for building ML-based infection risk predictors in this study, including experimenting across several data extraction and embedding configurations, architectural arrangements, and survival task prediction heads. Comparisons of the various experiments across standard performance outcome metrics and calibration statistics are presented in Table 2. Performance outcome metrics are computed over 100 bootstrap samples, with 1000 samples per bootstrap, as determined from sample size estimations. Manual feature extraction was performed across each individual patient’s timeline over variable look-back windows over 3–10 days were combined with static meta data, to create a multi-resolution summary of each patient’s health status. Additionally, a variational autoencoder was utilised to automatically extract data-driven features from 10-day windows across an individual patient’s timeline. Both manual and automatically learned feature embeddings were analysed for downstream survival task performance using traditional cox-proportional hazards (CoxPH) models (also known as DeepSurv [75] when parameterised with a deep network), a gradient boosted decision tree (GboostSurv) and time-discretised survival heads, LogHz and DeepHit.

**Table 2:** Risk stratification performance outcomes for hospital acquired infections. Comparison of calibration and performance outcome metrics between model risk predictors explored in this study and NEWS2 scores, the current standard of care. Performance outcome metrics are computed over bootstrap samples. ↑ indicates higher is better, ↓ indicates lower is better.

| | C-Index (↑) | AUROC (↑) | | | IBS (↓) |
| --- | --- | --- | --- | --- | --- |
|  |  | t=72 hrs. | t=24 hrs. | t=6 hrs. |  |
| NEWS2<sup>1</sup> | 0.604 (0.563–0.642) | 0.646 (0.632–0.667) | 0.734 (0.711–0.763) | 0.811 (0.769–0.862) | 1.726 (1.432–1.969) |
| CoxPH | 0.655 (0.621–0.713) | 0.724 (0.707–0.747) | 0.797 (0.771–0.823) | 0.865 (0.829–0.901) | 0.102 (0.094–0.109) |
| GboostSurv | 0.679 (0.632–0.718) | 0.730 (0.708–0.750) | 0.807 (0.777–0.835) | 0.898 (0.855–0.919) | 0.089 (0.081–0.097) |
| LogHz | 0.634 (0.608–0.670) | 0.660 (0.619–0.714) | 0.747 (0.673–0.806) | 0.747 (0.673–0.806) | 0.877 (0.865–0.886) |
| DeepHit | 0.634 (0.603–0.675) | 0.679 (0.630–0.732) | 0.769 (0.690–0.836) | 0.845 (0.770–0.948) | 0.131 (0.127–0.134) |
| VAE + CoxPH<sup>2</sup> | 0.743 (0.713–0.774) | 0.776 (0.741–0.811) | 0.796 (0.757–0.834) | 0.878 (0.829–0.914) | 0.177 (0.154–0.204) |
| VAE + GboostSurv<sup>3</sup> | 0.733 (0.713–0.751) | 0.765 (0.745–0.790) | 0.783 (0.756–0.804) | 0.825 (0.798–0.846) | 0.187 (0.184–0.190) |
| VAE + LogHz<sup>4</sup> | 0.750 (0.715–0.779) | 0.784 (0.747–0.822) | 0.801 (0.763–0.842) | 0.801 (0.763–0.842) | 0.745 (0.715–0.776) |
| VAE + DeepHit | 0.717 (0.683–0.745) | 0.758 (0.720–0.790) | 0.785 (0.743–0.819) | 0.856 (0.808–0.894) | 0.241 (0.240–0.242) |

<sup>1</sup> NEWS2, the national early warning score.
<sup>2</sup> VAE + CoxPH, also known as DeepSurv [75].
<sup>3</sup> VAE + GboostSurv, variational autoencoder embedding without the survival head loss back-propagated; post-hoc training of the GboostSurv with embeddings.
<sup>4</sup> LogHz, logistic-hazard, also known as Nnet-survival [61];
C-Index, concordance index, incorporating the inverse probability of censoring weights (IPCW) adjustment for highly censored data, at right-censoring cut-point, $t > 7$ days;
AUROC, time-dependent area under the receiver operating characteristic curve, at specified time-to-event, $t$ ;
IBS, integrated brier score, a measure of overall model calibration across all time points.

The VAE-derived feature embeddings demonstrated stronger predictive performance in estimating infection risk at longer time-to-event intervals compared to other feature extraction methods. However, as time-to-event intervals shortened and approached the onset of infection, the performance gap between VAE embeddings and manually extracted features narrowed, with all methods converging in predictive accuracy. Notably, within shorter intervals, the gradient-boosted survival model (GboostSurv) maintained competitive performance, even against more complex architectures.

Across all time-points, calibration and performance metrics of the machine learning-based survival models exceeded those of NEWS2, the current standard-of-care for detecting onset of infection in this healthcare setting. An exception was observed with the LogHz model, which despite showing a predictive power in rank-ordering patient risk over time, faced challenges in maintaining calibration consistency over time. Among the tested configurations, the VAE + CoxPH (DeepSurv) model exhibited the highest calibration accuracy across intervals, demonstrating particular robustness in cumulative risk prediction. GboostSurv also performed strongly across all metrics as time-to-event approached, making it a reliable and light-weight option for nearterm infection risk stratification. Both frameworks provided more reliable assessment of risk for identifying an infection event than NEWS2 scores, over a 7-day time horizon. For example, GboostSurv model estimated risk scores provided a more reliable assessment of risk for identifying an infection event than NEWS2 scores, as measured by a higher C-index for device estimated risk scores 0.679 (0.632–0.718) than NEWS2 scores, 0.604 (0.563–0.642), p<0.001. Fig. 2 presents the receiver operator curves for that framework, depicting the true positive and false negative rates for device risk scores in comparison to NEWS2 scores for discriminating various times to infection events. Comparatively, the VAE + CoxPH framework outperformed NEWS2 across all metrics, significantly achieving a higher C-Index, 0.743 (0.713–0.774) p<0.001, indicating stronger overall predictive accuracy. In terms of AUROC, VAE + CoxPH showed superior discrimination at each time interval, with scores of 0.776 at 72 hours, 0.796 at 24 hours, and 0.878 at 6 hours, compared to NEWS2’s respective scores of 0.646, 0.734, and 0.811, all p<0.0001. Additionally, the GboostSurv and VAE + CoxPH models had low integrated Brier scores (IBS, 0.089 and 0.177, respectively), reflecting improved calibration, and it demonstrated better alignment of predicted risks with actual outcomes over time.

**Figure 2:**
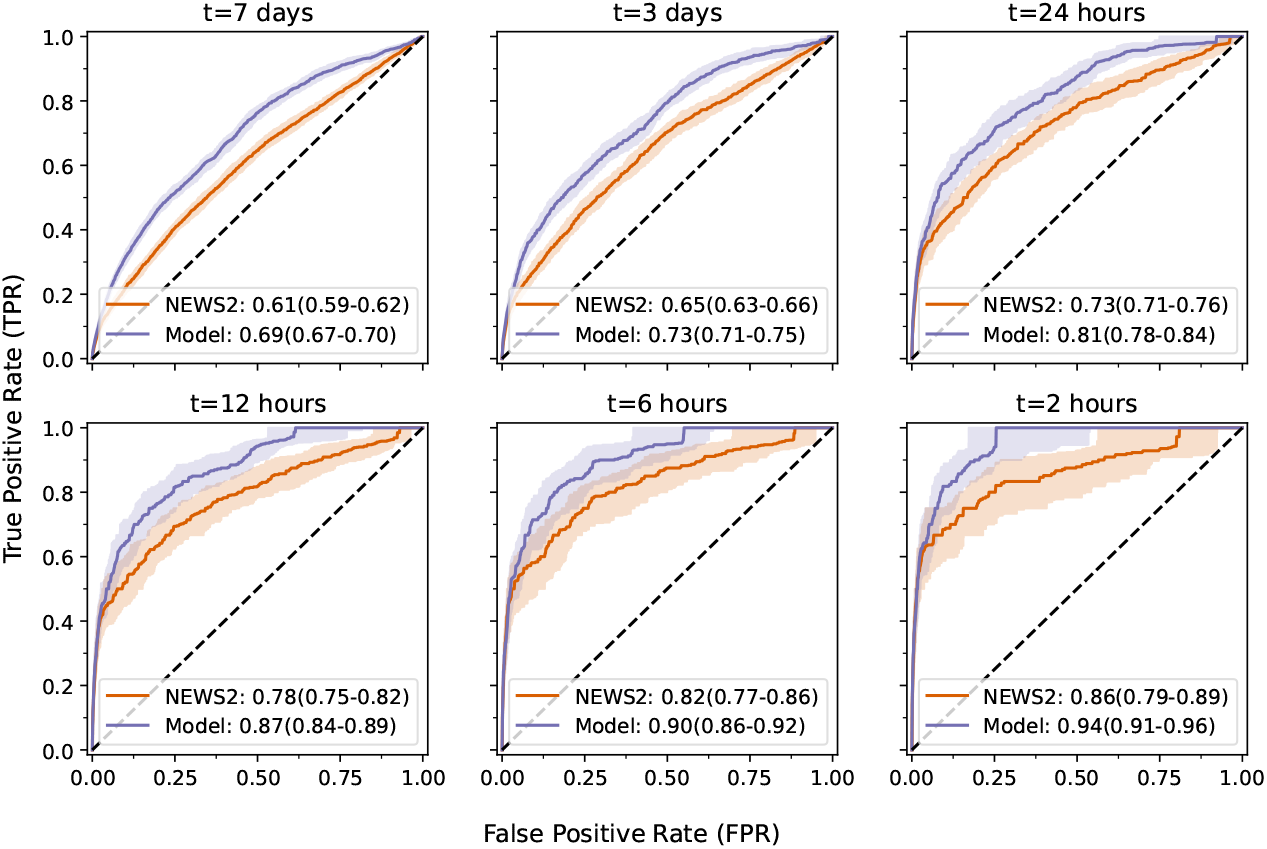
Analysis of model performance characteristics in comparison to NEWS2. Area under the receiver operator curves, *auroc*(*t*), at shortening times, *t*, to infection events for NEWS2 as compared to our model. Plots depict the median + IQR auroc (shaded-area) over 100 random bootstrap samples of the held-out data.

### 3.2 Stratification of patient risk levels in “silent-mode”

Model estimated risk scores were also visually assessed for effectiveness to stratify patients according to their infection susceptibility. Posterior risk scores were discretised into four equal-width bins based on the in-sample training set, defined as ordinal risk levels: low-risk, moderate-risk, high-risk, and critical-risk of suspected infection onset. Illustrative examples of the survival rates across risk-level groups in the out-of-sample testing set are presented in Fig. 3. Comparison of survival rates between model defined critical-, high-, and moderate-risk groups from low-risk and baseline survival rates was determined using log-rank tests. Baseline survival rates were calculated on the entire test set. It was found that the survival distributions between critical-, high-, moderate-risk groups were significantly lower than low-risk groups and from baseline survival rates in the control set, all p<0.0001. While it was observed that low-risk patient survival rates did not significantly differ from baseline survival (p=0.48), based on the required sample size (n≈25k samples) after adjusting for high censoring of low-risk and baseline-risk (censoring, 0.91–0.93), it was deemed that not enough patient measurements were available to determine statistical significance and thus the evaluation of low-risk against baseline risk was considered under-powered. All architectures investigated in the study demonstrated similar effectiveness to stratify patient risk levels, except for the LogHz model. The LogHz model exhibited poor calibration, as indicated by an observed flat survival prediction curve over time, failing to reflect the expected decline in survival probabilities as the event approached. This pattern suggested an inability to dynamically adjust risk in this use case, resulting in consistently misaligned predictions that do not accurately capture the hazard trajectory, as further reflected by its high IBS of 0.745.

**Figure 3:**
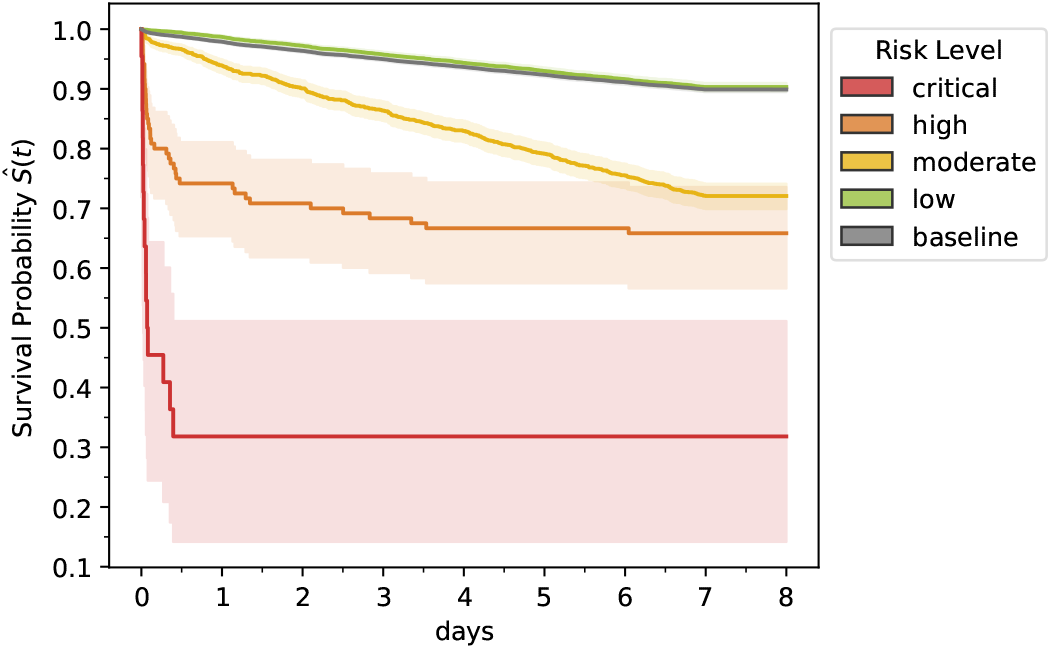
Silent-mode evaluation of risk stratification performance for hospital acquired infections (HAI). Evaluation of ML-derived risk prediction model in the out-of-sample test data. Predicted risk scores [log-hazard-ratio] were first grouped by [low, moderate, high, or severe]-risk of developing a HAI. Kaplan-Meier estimator of the survival functions for each group were empirically calculated over a 7-day horizon. Baseline survival rates were calculated over the held-out test data for comparison. A log-rank test indicated that survival rates for [moderate, high, and critical]-risk levels were significantly lower than the low-risk group and baseline survival rates (p<0.001).

### 3.3 Exploring interpretability of infection risk predictions

In order to enhance interpretability of the infection risk predictions, SHAP (SHapley Additive exPlanations) analysis was explored and applied to the manually extracted features within the GboostSurv model. SHAP analysis provided insights into feature importance by quantifying each feature’s contribution to individual risk predictions, making the model’s decision-making process more transparent. GboostSurv was chosen for SHAP analysis due to its strong predictive performance over shorter (near-event) lead times and its adaptability to complex data; its high performance with manually extracted features, which roughly matched the predictive power of VAE-derived features but were more interpretable, and combined with its tree-based structure allowed us to use SHAP to effectively explore model explainability.

We demonstrated how the model performance can be interpreted in Fig. 4 through SHAP based explanations. SHAP-based explainability uncovered the most relevant features for predicting HAI and survival. Top features included trends in patient histories, such as “days since last event” or the “number of previous events”; also current measurements, such as heart rate, as well as summaries over recent trends in vitals were useful. For example, the mean values or variance of temperature in the last 5– and 10–days. The trend (slope) of the heart rate, and demographic factor such as age, were also useful to explain survival times and outcomes. SHAP explainability also allows us to infer insights into the contribution of individual features. For instance, the days since last infection is highly predictive of future infections; the more recent the last infection (i.e. lower values), the more likely the patient will have another infection. Concurrently, if a patient has had fewer previous infections, often they will be less susceptible in the future. Other vitals-based examples include the mean temperature in the last 3–5 days. Higher maximum temperatures signify increased likelihood of upcoming infection.

**Figure 4:**
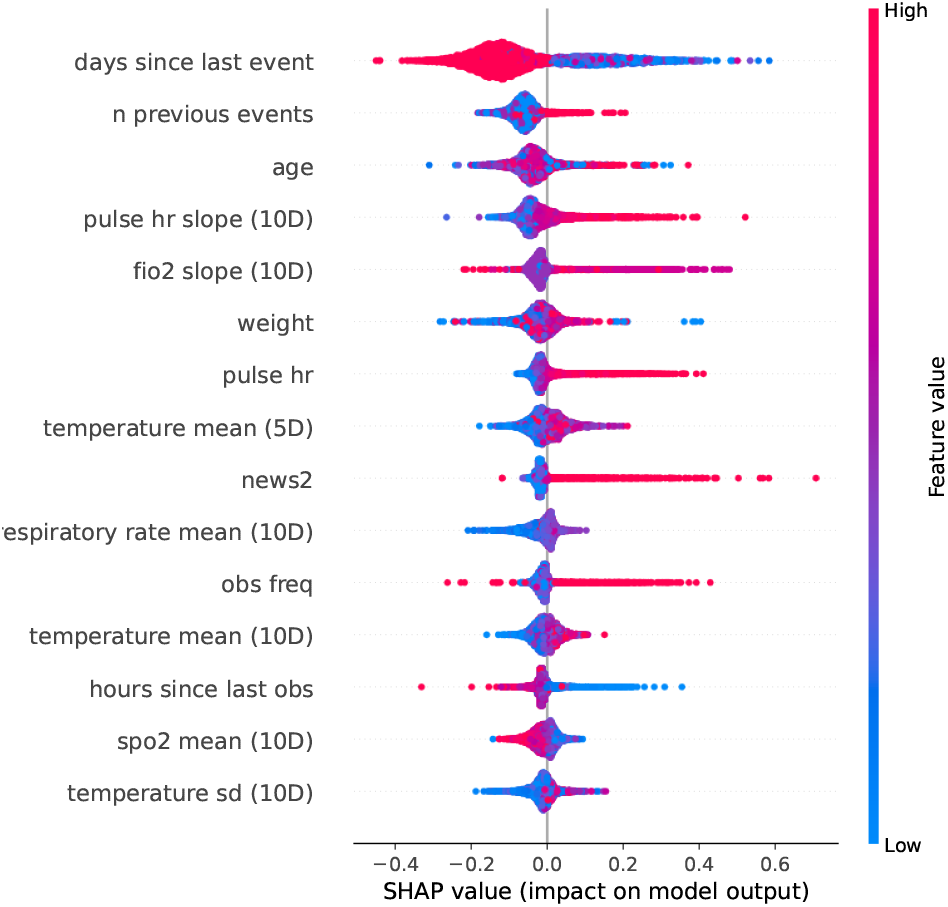
Global explainability of infection risk predictions. SHAP-based feature importance ranking for the top ranking features. Higher absolute feature importance, as measured by SHAP values, indicate a greater impact on the model’s prediction outcome; positive SHAP values indicate features that contribute to shorter survival times, while negative SHAP values represent features that are associated with longer survival times.

The combination of patient-specific model-estimated survival functions and augmented feature importance plot create a personalised explainability dashboard for each individual, as demonstrated in Fig. 5. As an example, we compared two randomly selected patients in the held-out set; one patient who experienced a HAI and one patient who was right censored at t=7 days. We compared the model-estimated survival predictions over the 7-day horizon as well as the accompanying feature importance plot for that observation set, as represented as local SHAP values. The patient in Fig. 5a is right-censored at t=7 days. The model predicts that there the probability that this patient will be event-free in 7 days is ≈0.80. Individual SHAP importance’s indicate that a low variance in this patient’s temperature and a lower maximum temperature in the last 10–days signified that this patient would not develop an infection, bolstered by the information that this patient has a lower number of previously recorded infections. In comparison, the patient in Fig. 5b developed an infection at t=1 days from model prediction (as indicated by a vertical line). In this example, the model predicted that the likelihood this patient will be event-free after 7 days was close to 0. At the time-of-event, the model estimated a ≈0.6 chance of developing a HAI.

**Figure 5:**
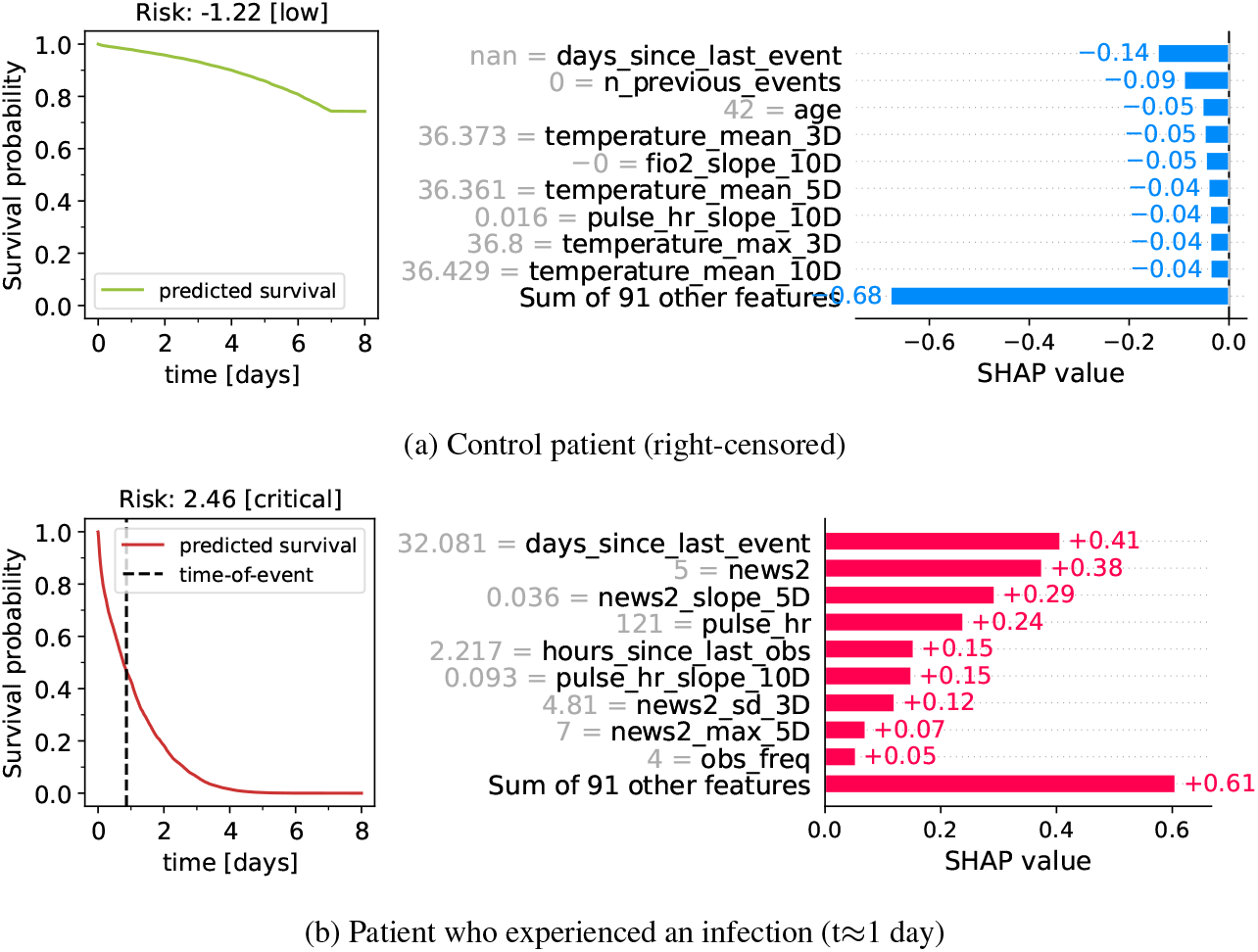
Personalised explainability for individual patients. Comparison of model-estimated survival predictions for two example patients in the held-out test set: **(a)** a patient who was right-censored at t=7 days and **(b)** a patient who developed an infection at t ≈1 days from model prediction. The actual time-of-event is noted by a vertical line, otherwise the data is considered right-censored. The accompanying feature importance plot, represented as local SHAP values, corresponds to the same model-estimated prediction for that individual. Higher feature importance indicates a greater impact on the model’s prediction outcome; positive SHAP values indicate features that contribute to a shorter survival time, while negative SHAP values represent features that are associated with longer survival times for these individuals. Note, ‘nan’ days since last event indicates that this patient has not experienced a previous infection in their historic dataset.

## 4 Discussion

In this study we developed a machine learning framework aimed at identifying elevated risk of infection for patients with complex neurological impairments continuously over a 7-day horizon. We demonstrated that data collected from historical multimodal patient meta-data, combined with recent trends in observational biomarkers, such as vital signs, clinical reported outcomes, and information extracted from patient clinical notes, could be utilised to robustly estimate the likelihood and timing of clinical suspicion of infection, as measured by first antibiotic administration. Further, by categorising model-generated risk scores as [low, moderate, high, severe]-risk via data-driven methods, we could effectively stratify patients according to their infection susceptibility. We explored manual and data-driven (generative deep learning) methodologies to extract rich individual patient-level representations across multiple experiments testing various survival head estimators (CoxPH, GboostSurv, LogHz, and DeepHit). We found that the GboostSurv and VAE + CoxPH configurations consistently outperformed others at identifying infections (AUROC, *>*0.878 up to 6 hours prior to an antibiotic prescription) and exhibited excellent calibration (IBS, <0.178), indicating that predicted infection risks closely matched the actual observed infection rates over time. In most instances, model estimated scores provided more reliable risk assessments identifying clinical suspicion of infections than the current standard of care in the healthcare setting examined, the NEWS2 scoring system. For instance, model estimated risk scores achieved a higher C-index for device estimated risk scores than NEWS2 scores and had higher discriminative ability to correctly distinguish between individuals who experienced the event and those who do not at specific time points as measured by a time-dependent AUROC in comparison to NEWS2, at all t ≥6 hrs. to an event. As such, our findings support the suitability of ML-based systems to stratify infection risk in real-world hospital settings. Finally, our retrospective 1–year silent mode evaluation allowed us to observe how predicted risk levels aligned with actual survival outcomes, enabling precise calibration and validation of the model’s predictions in a controlled setting without impacting patient care, in preparation for forthcoming real-world deployment.

This study’s observations align with existing research that employs machine learning for infection risk prediction and on similar tasks of interest, yet we introduce notable distinctions in methodology and outcomes.

While direct comparisons are difficult, broadly we achieved a similar performance to other machine learning studies predicting nosocomial infections, for instance Feng et al. (AUROC, 0.85 t*>*48 hrs.) [18] and Chen et al. [76] (AUROC, 0.750–0.834), however the latter study is based on monthly cross-sectional surveillance data, rather than granular electronic health record (EHR) data. Notably, in the model developed by Feng et al.—also exploring a gradient boosting ensemble of decision trees, XGBoost—laboratory measurements appear as top predictive features, however, are not currently available to our model. Given we achieved a similar AUROC levels without additional laboratory data, integrating these data sources into our framework could potentially yield significant improvements in downstream performance. Our Conditional Variational Autoencoder (VAE) configuration demonstrated strong prognostic accuracy predicting HAIs (AUROC, 0.878 t≥6 hrs.), with the strongest comparative performance at earlier time-points (AUROC, 0.796 t≥24 hrs.; 0.776 t≥72 hrs.), potentially providing a greater “window of opportunity” for clinical teams before an infection event than other configurations. The relative performance of our VAE configuration is comparable to the few other studies that use state-of-the-art deep learning techniques for infection prediction [20], [77], one interesting example being the Double Fusion Sepsis Predictor (DFSP) by Duan et al. [77], which combines deep and handcrafted features for early sepsis prediction. Interestingly, our GboostSurv model, utilising handcrafted features inspired by Hyland et al. [42], also exhibited competitive performance to the VAE + CoxPH configuration developed, particularity at short lead times to antibiotic prescription (AUROC 0.889 t ≥6 hrs.) This finding aligns with Hyland et al.’s work [42], which that “careful feature design” can achieve comparable performance to deep learning based approached for clinical prediction tasks. Moreover, the interpretability of the GboostSurv model is a significant advantage, as highlighted by Feng et al. [18], who underscore the importance of model transparency in infection risk prediction. Van der Vegt et al. [28], [29] further discuss the critical role of interpretability in ML clinical prediction models, particularly concerning real-world deployment of clinical decision support tools and clinician or user trust.

Our study was framed as a time-series survival task to capture not only the probability of healthcare associated infections but also the estimated timing of infection onset, which is crucial to enable early intervention strategies. Most studies building ML solutions to determine clinical deterioration—whether generally, or for specific instances like infections or sepsis—more often will take the form of time-series classification tasks rather than the survival-based tasks investigated in this work [13], [29]. Henry et al., however, the first to use

ML and EHR data to develop a scoring system (i.e., ‘TREWscore’) that predicted septic shock hours before onset utilised a CoxPH model (i.e. survival task) model to identify a subset of features most indicative of septic shock and generated a risk prediction score over time [30], [78]. Unlike a time-series classification approach, which simply predicts infection occurrence, survival analysis can also account for censored patients (those discharged or uninfected within the observation period), reducing bias in risk estimation and a flexibility to incorporate other tasks and confounders in future work. Furthermore, survival analysis offers insights into *when* an infection is likely to occur within a time-horizon, not just *if* and infection will occur across some fixed time-horizon. This timing information supports proactive, tiered interventions, and a “window of opportunity” allowing clinicians to prioritise patients based on imminent risk. Categorising risk scores into risk levels could further help facilitate real-world use of the model by providing clinicians with clear, actionable thresholds that support prioritisation of patients based on infection risk severity, enabling targeted interventions for those at highest risk, while allowing low-risk patients to be monitored with fewer resources. This temporal insight is also particularly valuable in infection control and antimicrobial stewardship, where earlier identification and diagnosis of infections will be critical for timely administration of the most appropriate antimicrobial drugs [14], and where early interventions can significantly reduce the likelihood of severe outcomes [79] and prevent infection spread [80].

In clinical AI, achieving a balance between model performance and explainability is essential, as highly accurate models may lack the transparency required for clinical adoption [81]. Although the VAE + CoxPH model demonstrated the best overall combination of performance and calibration at short and long lead times, the competitive results of GboostSurv close to an infection event, with AUROC values up to 0.889 t≥6 hrs. and a strong C-Index of 0.679, led us to favour GboostSurv for its inherent explainability. This emphasis on interpretability is crucial for clinical use, supports better adoption in clinical settings, where clear insights into risk factors and decision rationales are essential for integrating the model into patient care workflows [82]. Our analysis using SHAP to create a personalised explainability dashboard provided several key insights. First, it offered added interpretability by clearly visualising model feature importances both globally and for individual patients. Second, the analysis demonstrated that the model was performing as expected, as the top features identified by the model aligned well with clinical knowledge regarding infection identification. For example, trends in vital signs, such as temperature and heart rate—signs commonly indicative of infection— were predictive of infection onset. A patient’s historic susceptibility to infections, their age, and any recent infections were also predictors of new infections. The use of SHAP enabled us to understand the influence of features on individual patients, creating a personalised and explainable picture of each prediction that clinicians can use to tailor interventions. By combining SHAP-based insights with individual survival predictions, clinicians may gain a clear understanding not only of each patient’s risk level but also of the specific factors driving that risk.

While this study demonstrates promising results, several limitations must be acknowledged. First, the data used for training and validation were sourced from a single setting, which may limit the model’s generalisability across diverse patient populations, healthcare environments, and clinical practices. This specificity introduces potential biases in the patient population, including demographic and health condition distributions, which may not extend to broader settings. Additionally, differences in diagnostic equipment or procedural variations could affect the model’s applicability in other hospitals. The relatively narrow patient population highlights the need to expand the model’s validation to more diverse populations and healthcare environments. Another limitation involves the selection of the 7-day time horizon for right-censoring; a more optimised configuration, potentially including shorter or longer intervals, could yield more accurate risk assessments across varying patient trajectories. Temporal bias also presents a challenge, as evolving infection control practices and shifts in diagnostic criteria over time may have impacted infection rates and influenced model performance. Furthermore, the model’s performance relies on the quality and completeness of electronic health record data, which can vary widely across institutions. Finally, while GboostSurv was selected for its interpretability, this choice may trade off some predictive power that more complex models could achieve, and its lack of external validation leaves uncertainty about its robustness in new settings. Addressing these limitations through external validation, model recalibration, and expanded testing will be essential for assessing the broader clinical utility of this approach

Future research will focus on expanding the data modalities included in the model: lab data such as bloods, microbiology, pathology will be incorporated, as well as more advanced methodologies to further process the data already collected. For example, the integration of clinical language models to process unstructured clinical notes offers a promising avenue for extracting rich, contextual features that go beyond manual feature extraction, enhancing the model’s ability to capture nuances in patient health trajectories [83]. Importantly, our modular framework allows for easy incorporation of these new data types, ensuring the adaptability of the approach as healthcare data sources evolve. Further exploration of alternative VAE architectures, including those leveraging transformer-based survival models, could also enhance model performance by capturing more complex temporal dependencies in patient data. To improve generalisability, future studies will validate and calibrate our model configurations across multiple healthcare institutions, capturing more diverse patient populations, equipment, and clinical practices to ensure robust performance in varied settings. Although this study evaluated the model in ‘silent mode’ deployment, future work will collect outcomes from prospective, real-time implementation to assess its clinical utility and impact on patient outcomes in a live hospital setting. Expanding on these areas will not only address current limitations but will also drive meaningful advancements in infection risk prediction for diverse healthcare environments.

This study highlights the potential of machine learning models, incorporating both feature-based and deep generative machine learning approaches, to accurately predict infection risk and support proactive patient management. By balancing predictive strength with interpretability for real-world applications, these approaches demonstrate how AI-driven risk stratification can play a pivotal role in enhancing proactive clinical decision-making for patients with complex neurological impairments, aiding infection control and prescribing decision support, improving patient outcomes, and optimising resource allocation within strained healthcare settings.

## Data Availability

All data referenced in this manuscript are hospital data and are therefore not publicly available due to patient privacy and confidentiality restrictions. Access may be granted on request to the corresponding author, subject to relevant institutional and ethical approvals.

## Acknowledgements

The authors thank all study participants and their families for their time and contributions to this research. We also acknowledge the staff at the Royal Hospital for Neuro-disability for their support and collaboration, especially Rebekah Van Slyke, Toby Roberts, and the Information Technology team, as well as PatientSource, for their assistance in facilitating this work. Our thanks also go to Eddie Pease, Richard Silk, Anastassia Rowe, Tiia Meuronen, Nita Shah, and Waquid Valiya Peedikakkal for their contributions to the work presented in this study.

## Disclosures

A.P.C is an employee and shareholder at Sanome, UK. T.P. is a shareholder at Sanome and was employed at Sanome during the completion of this work. P.A is an employee and a shareholder of PatientSource. L.B is an employee of RHN. S.D. is an employee of RHN.

## A Appendix

**Figure A.1:**
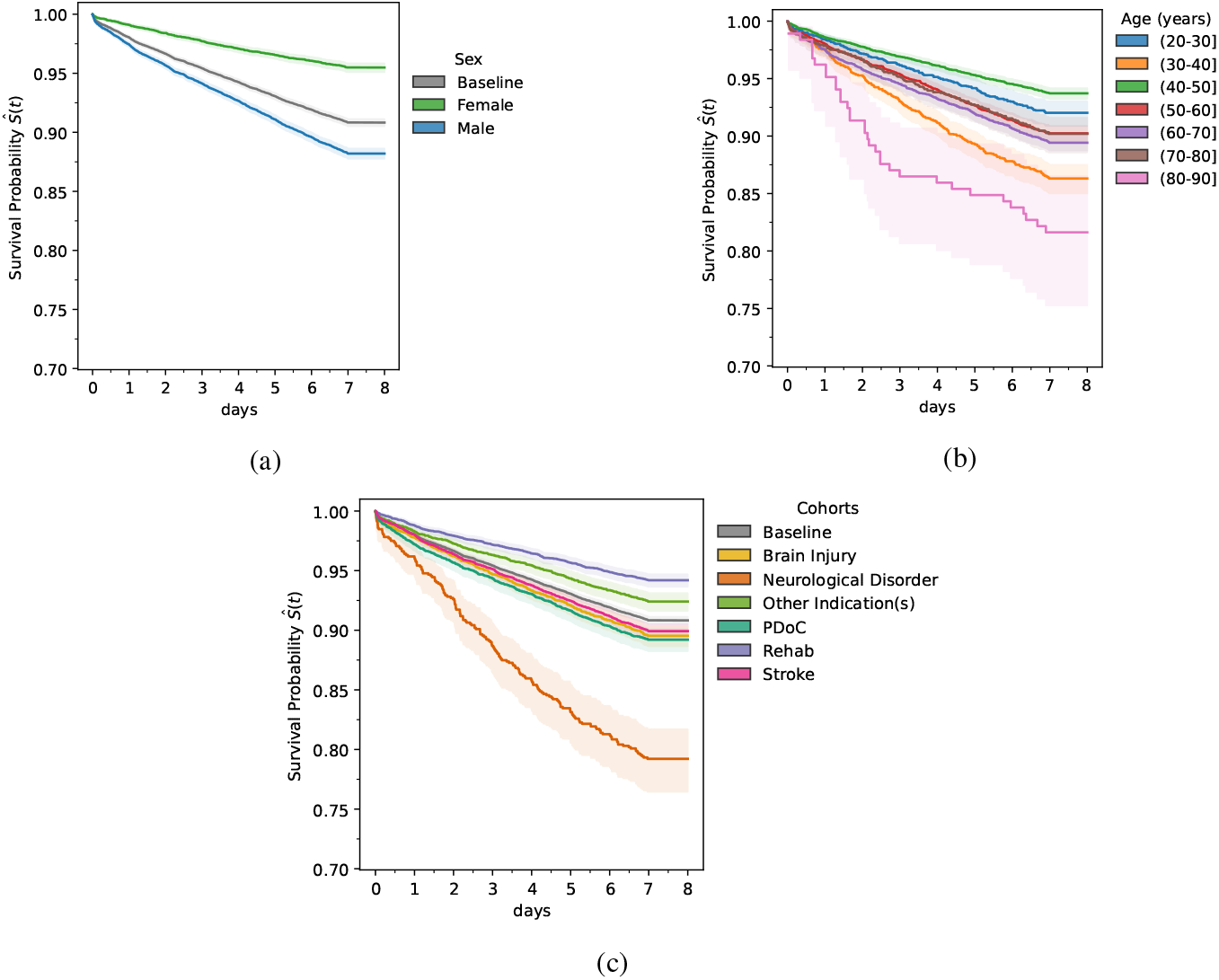
Infection survival rates by demographic factors and comorbidities. Empirically calculated Kaplan-Meier estimator of the survival functions by (a) sex; (b) 10-year age range groups for 7-day infection estimation. Baseline survival rates are calculated over the entire study population; (c) major co-morbidity subgroups: brain injury, neurological disorders, other conditions, prolonged disorders of consciousness (PDoC), rehab, and stroke patients.

## Notes

### Funding Statement

This study was funded by Sanome Ltd.

### Author Declarations

This study was conducted in accordance with the ethical principles outlined in the Declaration of Helsinki and received favourable ethical opinion from NHS Research Ethics Committee 24/NE/0008 supported by the Confidentiality Advisory Group 23/CAG/0110. All data handling and analysis were performed in compliance with confidentiality guidelines and data protection regulations. The use of retrospective patient data was ethically reviewed, ensuring the study adhered to principles of transparency, data privacy, and minimal risk to participants.

